# Deep learning-based stride segmentation with wearable sensors: effects of data quantity, sensor location, and task

**DOI:** 10.1101/2024.12.17.24319160

**Authors:** Anthony J. Anderson, Michael Gonzalez, David Eguren, Naima Khan, Isabella Zuccaroli, Siegfried Hirczy, Valerie E. Kelly, Brittney C. Muir, Kimberly Kontson

**Affiliations:** US Food and Drug Administration, Center for Devices and Radiological Health, Office of Science and Engineering Laboratories, Silver Spring, MD, USA; University of Washington, Department of Mechanical Engineering, Seattle, WA, USA; University of Washington, Department of Neurology, Seattle, WA; VA Puget Sound Health Care System, Seattle, WA, USA; University of Washington, Department of Rehabilitation Medicine, Seattle, WA, USA

## Abstract

Accurate stride segmentation from wearable sensors is foundational for digital gait assessment tools, yet systematic evaluations of deep learning approaches across varied real-world mobility tasks remain limited. We developed and assessed Temporal Convolutional Network (TCN) models for stride segmentation using data from 121 older adults with and without Parkinson’s disease, specifically evaluating how performance varies with model development data quantity, sensor location, and movement complexity. Using a fixed-size test set of 40 participants, we found that lower limb sensors achieved F1 scores above 95% during walking with just 5–10 training participants, but performance declined substantially during more complex movements such as 180° turns. Foot-mounted sensors maintained robust performance across tasks (F1: 99.3% walking, 96.7% turning, 88.4% stationary and transitional movements), while wrist sensors showed marked degradation (F1: 88.4% walking, 72.3% turning, 50.0% stationary and transitional movements). Our findings demonstrate that it is important to tailor performance testing for digital gait assessment tools to both sensor location and use case, as performance achieved during controlled walking may not generalize to complex movements in daily life.

## Introduction

Digital health technologies that incorporate wearable movement sensors hold the potential to transform our ability to diagnose mobility-limiting disorders, identify prognostic indicators, and assess therapeutic interventions^1–5^. Inertial measurement units (IMUs) are attractive tools for gait analysis, enabling the collection of continuous movement data in various environments. One common approach for wearables-based gait analysis involves deriving statistical descriptions of spatiotemporal gait metrics, such as stride length and gait speed^6–9^. However, before these metrics can be computed, a crucial preprocessing step is required: the segmentation of individual strides from a collected bout of movement via the identification of gait events (i.e., initial contact). This segmentation process serves as the algorithmic backbone for wearables-based gait assessment, as all downstream spatial and temporal metrics depend on the accurate identification of stride boundaries in time.

Many methods for automatically segmenting strides from IMU waveforms have been presented in the literature, with a focus on algorithms developed for sensors placed on the foot^10–17^, shank^18–21^, and lower back^21–23^, and less emphasis on other sensor locations^24^. While numerous studies report high segmentation accuracies, the generalizability of model performance is often confounded by locomotor task complexity. Approaches like peak detection and Multidimensional Subsequence Dynamic Time Warping (msDTW), which rely on expected waveform morphologies, have shown success in controlled, straight-line walking scenarios^10,18,19^ but tend to struggle in more complex settings, e.g., 180-degree turns^13,19^. In contrast, machine learning algorithms that learn to identify stride boundaries directly from data have shown promise in handling more diverse and challenging pathological movement patterns^13^. Recent advancements in deep learning, particularly Temporal Convolutional Networks (TCNs)^25^, have demonstrated high performance in gait tasks^16,20,21,24^, and potentially offer a more robust solution for stride segmentation across various conditions.

TCN models, and deep learning approaches in general, have strong potential as a paradigm for stride segmentation, addressing several limitations of traditional methods. Unlike heuristic approaches that rely on predefined rules regarding expected waveform shapes or methods that rely on *a priori* template signals, deep learning models are trained to identify gait events directly from data. This data-driven approach allows for automatic extensibility to various types of gait events present in the training dataset, including complex movements like 180-degree turns. Deep learning methods can potentially capture a variety of noncyclic movements in the data that may be missed by traditional algorithms. Additionally, the model development process is independent of sensor location, providing flexibility in deployment across different body sites^24^. However, these advantages require the collection of training data with reliable ground truth labels, which can be expensive and time consuming.

Despite the promising advantages of TCN approaches for stride segmentation, several important gaps remain in our understanding of their performance across various dimensions. The quantity of training data required to achieve stable and generalizable segmentation performance is not well established, which leads to uncertainty in study planning and algorithm development timelines. Additionally, the impact of sensor location has not been systematically evaluated in pathological cohorts, limiting our ability to optimize sensor placement for different clinical applications. Furthermore, most existing studies have focused on algorithm performance during straight-line walking^10,11,18,22^, leaving open questions about how performance might change when assessing more complex movements such as turning. These knowledge gaps hinder the development and implementation of rigorous ‘analytical validation’ studies, as defined in the Digital Medicine Society’s V3+ framework^26^. Addressing these gaps is important for advancing the field of wearable sensor-based gait analysis and realizing the full potential of deep learning approaches in clinical and research applications.

To address these gaps, we present a detailed analysis of TCN model performance for stride segmentation across multiple training data volumes, sensor locations, and movement tasks using the WearGait-PD dataset, a large open-access movement dataset of older adults and people with Parkinson’s disease (PD)^27^. Our novel contributions include: (1) an evaluation of model performance as new participants were added to the training and validation datasets for a fixed size test dataset, (2) a description of how these results vary across eight unique sensor locations, and (3) a sub-analysis of how the results vary between straight-line walking, turns, and other complex movement tasks. We hypothesized that (H1) average model performance on the unseen test set would increase as participants were added to the training set, (H2) sensors placed on the lower body would demonstrate higher performance than sensors placed on the lower back or upper body, and (H3) model performance would be higher during straight-line walking than during turns and other tasks. Figure 1 shows an overview of the study.

**Figure 1:**
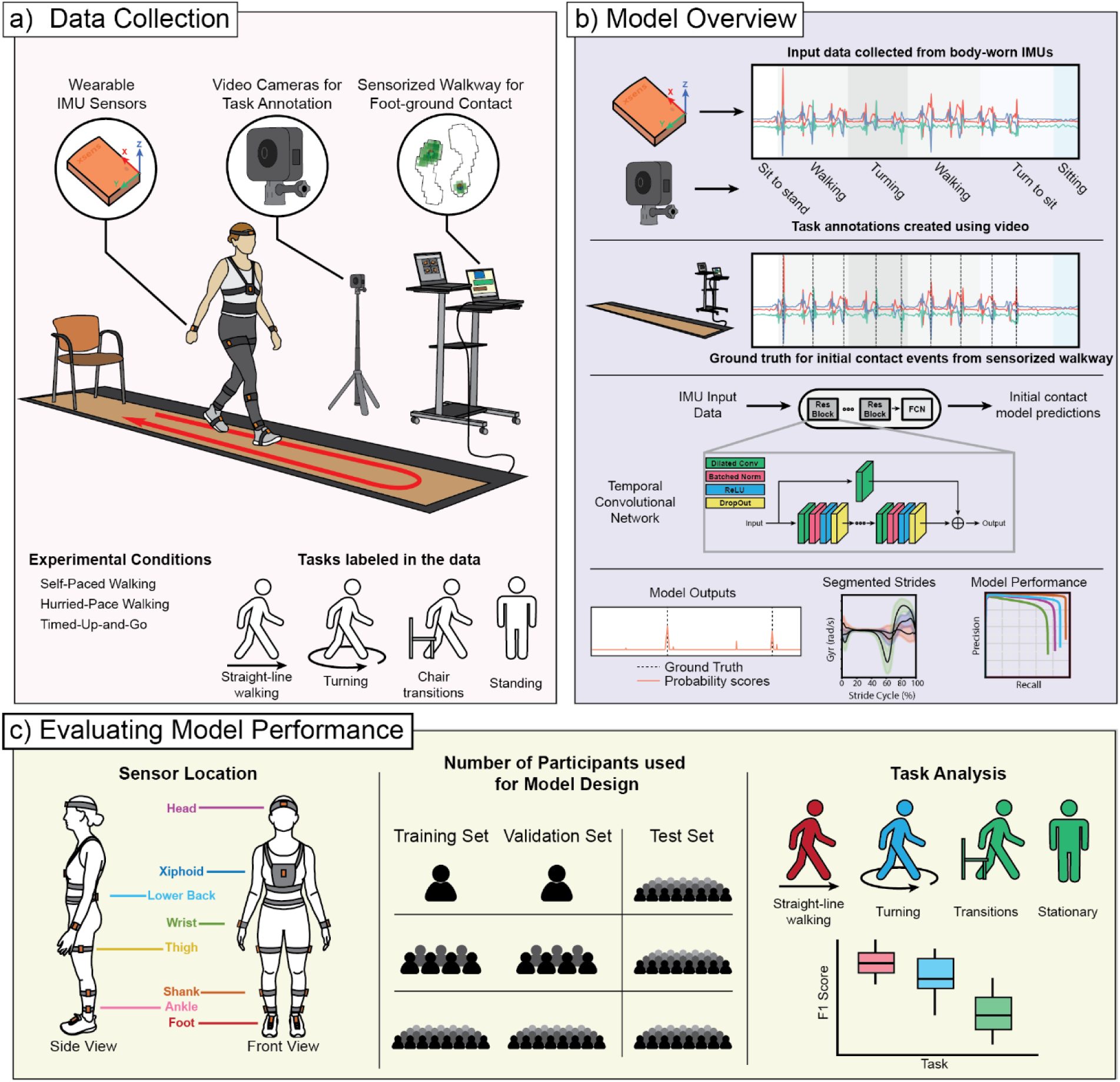
Overview of the study methodology for evaluating stride segmentation using wearable sensors. a) Data collection setup showing a participant wearing eight inertial measurement units performing movement tasks on an instrumented walkway. Experimental conditions included self-paced and hurried-pace walking, with and without 180-degree turns on the walkway, and the Timed Up and Go Test. Movement tasks were labeled through video annotation. b) Model development pipeline showing representative IMU accelerometry signals with corresponding video-based task annotation and walkway-derived ground truth stride boundaries, followed by the TCN architecture used for stride segmentation and example model outputs. c) Study design exploring how model performance is impacted by three key dimensions: sensor location (eight anatomical positions), training data quantity (evaluating model performance with increasing participant counts while maintaining a fixed-size test set), and movement task complexity (walking, turns, and stationary and transitional movements).

## Results

We evaluated the performance of TCN models for initial contact detection across multiple sensor locations and training data quantities using data from 121 participants. Our analysis framework incorporated a 10-fold cross validation approach, with each fold maintaining distinct training, validation, and test sets. The number of initial contact events assessed varied by sensor location due to wireless signal dropout (see Supplementary Table S1 for exact counts), with a mean of 26,693 initial contact events analyzed across the full dataset and a mean of 222 events per participant. Model performance was evaluated using precision, recall, and F1 scores. We systematically investigated how model performance varied with increasing training data quantity (2-81 participants split equally between training and validation sets), sensor location (8 distinct body locations), and movement complexity (walking, turning, and stationary and transitional movements), while evaluating the model on a fixed-size test set of 40 participants. Performance varied substantially across these dimensions, with mean F1 scores ranging from 99.3% [99.1, 99.5, 95% CI] for the dorsal foot sensor during walking to 50.0% [48.5, 51.7] for the wrist sensor during stationary and transitional movements when trained with the maximum dataset size. Additionally, we examined model performance between PD and control participants to verify the robustness of our findings across both groups.

### Participant Demographics and Clinical Scores

Study data were drawn from a large open-access dataset^27^ collected across two geographic locations, Seattle, WA and Baltimore, MD. At the time of this analysis, the dataset contained 121 participants: 58 individuals with PD (mean age 65.9 ± 8.8 years, 23 female) and 63 controls (mean age 76.3 ± 8.5 years, 39 female). Among PD participants, the mean MDS-UPDRS Part III motor score was 24.5 ± 10.3, and the median Hoehn & Yahr stage was 2 (minimum 1, maximum 3), indicating that most participants had bilateral symptoms with minimal to moderate disability but maintained independent physical functioning. Detailed participant characteristics are provided in Table 1.

**Table 1:**
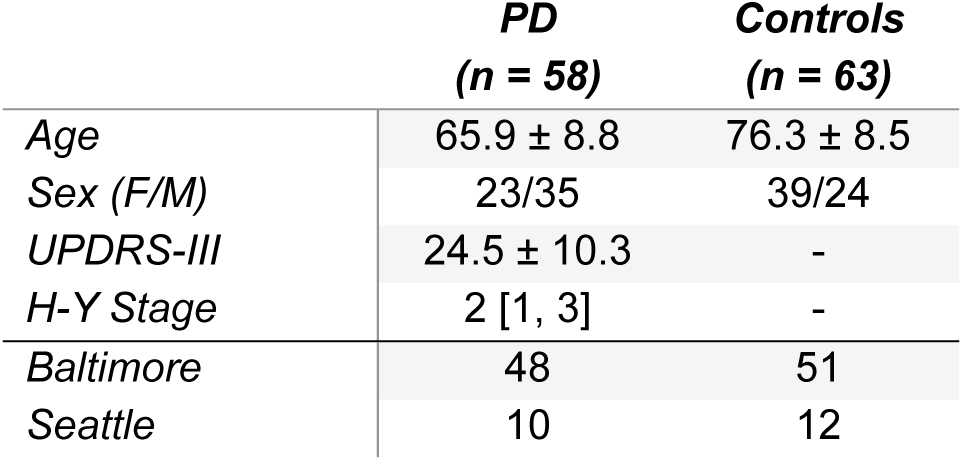
Participant demographics, clinical scores, and data collection locations. Mean and standard deviations values are presented, except for H-Y Stage, which is shown as median [min, max].

### Effects of Training Data Quantity, Sensor Location, and Task

We examined how TCN model performance for initial contact detection varied with training data quantity, sensor location, and movement complexity. Figure 2 illustrates these relationships across eight sensor locations, showing mean F1 scores and 95% confidence intervals from our cross-validation analysis. Model performance exhibited characteristic learning curves across all sensors and movement types, with rapid initial improvement as training and validation participants were added, followed by more gradual gains or plateaus. Performance patterns differed markedly between movement types, with consistently higher F1 scores during walking tasks compared to turning or stationary and transitional movements. This effect was particularly pronounced for sensors placed farther from the feet. Notably, while all lower-limb sensors achieved high performance (F1 > 90.0%) for walking tasks with sufficient training data, performance degraded during complex movements, especially for sensors placed on the upper extremity.

**Figure 2:**
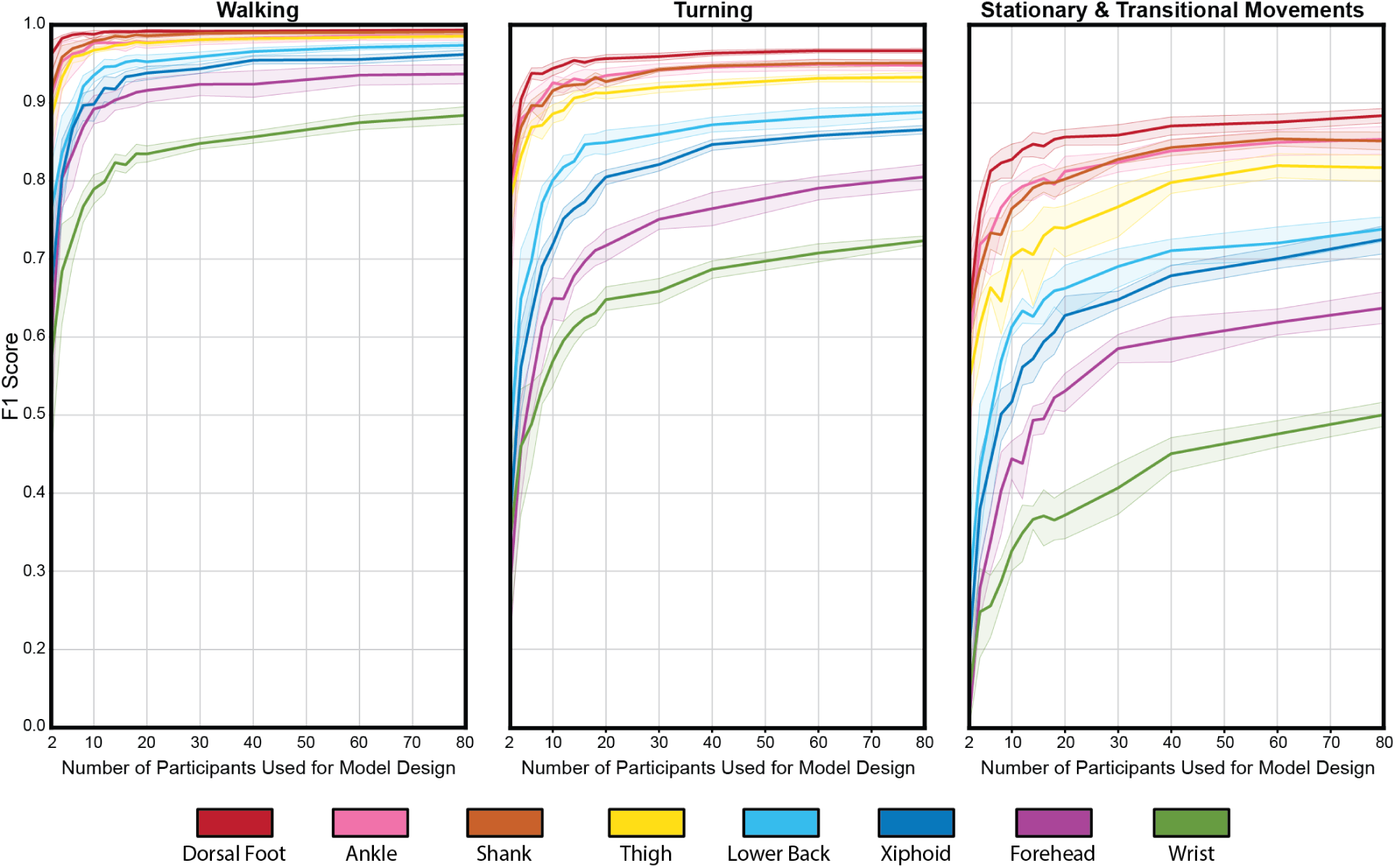
Model performance across sensor locations, movement types, and training data quantity. F1 scores (mean and 95% confidence intervals from 10-fold cross-validation test sets) are shown as a function of the number of participants used for model design (training and validation sets) during walking, turning, and stationary and transitional movements. For each cross-validation fold, the test set uniformly contained 40 unseen participants.

### Data Quantity

The relationship between model performance and training data quantity exhibited distinct patterns across sensor locations and movement types (Figure 2). During walking tasks, lower-limb sensors (dorsal foot, ankle, shank, and thigh) showed rapid initial improvement in F1 scores with relatively small datasets. For these sensors, performance tended to taper or plateau in performance between 2 and 10 participants. In contrast, sensors placed on the trunk and head required more training data, with performance tapering occurring between 10 and 20 participants. Across all participant counts, the wrist sensor had relatively low performance. However, its F1 score had a notably positive slope at the 80-participant mark, indicating that a larger dataset would likely lead to continued improvement.

These learning curve characteristics shifted during more complex movements. During turning tasks, while lower-limb sensors maintained similar learning curve shapes, they plateaued at lower F1 scores compared to walking. Trunk and head-mounted sensors not only showed decreased performance, but also required more training data to improve, with less distinct plateaus. This pattern was further amplified during stationary and transitional movements, where all sensors showed both lower absolute performance and more gradual improvement with increased training data.

### Sensor Location

Sensor location significantly impacted model performance (p < 0.001) when evaluated using the maximum training/validation dataset of 81 participants (Table 2) by a Friedman test (n = 10 cross validation folds for each of 8 sensors). A clear hierarchical pattern emerged across all tasks, with sensors placed on the lower extremity consistently outperforming those placed on the trunk, head, and upper body. The dorsal foot sensor achieved the highest performance, followed closely by the ankle and shank sensors, which performed similarly to each other. Performance generally decreased as sensor location moved superiorly, with the thigh sensor showing intermediate performance, followed by progressively lower performance for the lower back, xiphoid, forehead, and wrist sensors (Figure 2). A post-hoc Nemenyi test (n = 10 cross validation folds) for pairwise comparisons showed that the dorsal foot sensor significantly outperformed all sensors except the ankle, shank, and thigh (p < 0.001; see Table S2 for complete statistical comparisons). Conversely, the wrist performed significantly worse than all lower body sensors (p < 0.001). See Table S2 in the Supplementary Materials for all pairwise statistical comparisons between sensors.

**Table 2:** Mean test set F1 score and 95% confidence intervals across cross-validation folds for each sensor location and task. Each comparison was performed when 81 participants were used for model development. All values are expressed as percentages.

| *Sensor* | *Walking* | *Turning* | *Stationary & Transitional* |
| --- | --- | --- | --- |
| *Dorsal Foot* | 99.3 [99.1, 99.5] | 96.7 [96.4, 96.9] | 88.4 [87.5, 89.3] |
| *Shank* | 99.1 [98.8, 99.3] | 95.1 [94.7, 95.5] | 85.1 [84.0, 86.3] |
| *Ankle* | 98.6 [98.0, 99.1] | 94.8 [94.1, 95.4] | 85.3 [83.4, 87.0] |
| *Thigh* | 98.6 [98.3, 98.8] | 93.4 [92.8, 93.9] | 81.7 [79.9, 83.3] |
| *Lower Back* | 97.4 [97.0, 97.8] | 88.8 [88.0, 89.6] | 73.8 [72.2, 75.4] |
| *Xiphoid* | 96.2 [95.7, 96.8] | 86.6 [86.0, 87.1] | 72.5 [70.7, 74.2] |
| *Forehead* | 93.7 [92.5, 94.9] | 80.5 [78.9, 81.1] | 63.7 [61.7, 65.8] |
| *Wrist* | 88.4 [87.3, 89.5] | 72.3 [71.7, 73.0] | 50.0 [48.5, 51.7] |

The precision-recall curves (Figure 3) provide additional insight into model performance across sensor locations, demonstrating the achievable precision and recall values at different initial contact detection thresholds. These curves show that while high precision and recall are simultaneously achievable for lower limb sensors, there is a more distinct trade-off for trunk and upper body sensors. This analysis provides practical guidance for applications where either precision or recall might be prioritized over F1 score.

**Figure 3:**
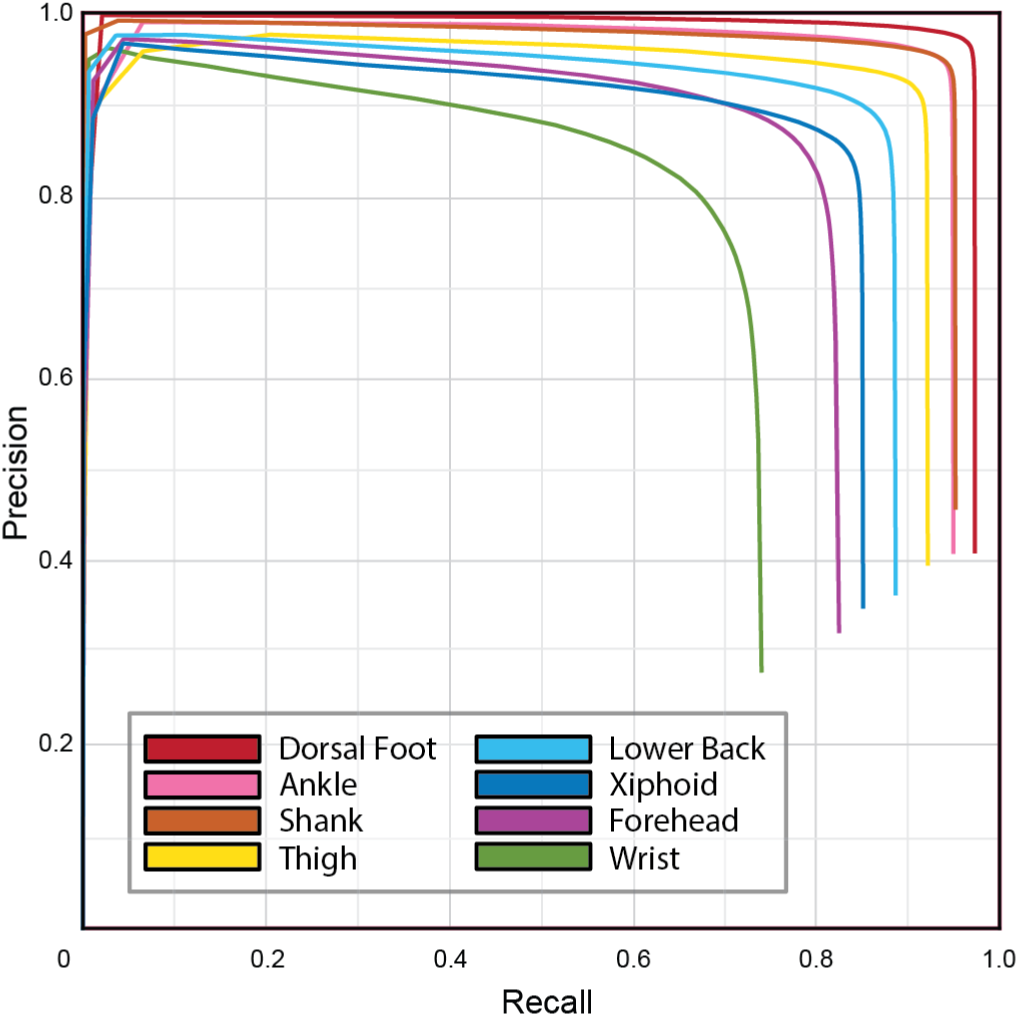
Precision-recall curves for stride detection across sensor locations for all movement tasks. Each curve represents the mean test set performance across 10 cross validation folds using models developed with the maximum model development dataset size of 81 participants.

### Task Complexity

Task complexity had a significant impact (p < 0.001) on model performance when averaging across all sensor locations and cross validation folds (Figure 2) as evaluated using the Friedman test (n = 80 cross validation fold × sensor combinations). Performance was highest during walking tasks, which included steady-state walking as well as gait initiation and termination, across all sensors (F1 scores 88.4%–99.3%) with the lowest score at the wrist. A post-hoc Nemenyi test (n = 80 cross validation fold × sensor combinations) for pairwise comparisons showed that performance decreased significantly (p < 0.001) during turning (F1 scores 72.3%–96.7%) compared to walking, though all lower limb sensors maintained F1 scores above 90% even during these more complex movements. Model performance for the stationary and transitional movements, including sitting, standing, and transitions into and out of a chair, ranged widely (F1 scores 50.0%–88.4%) where no sensor achieved an F1 score above 90%. Model performance on these movements was significantly lower compared to both walking (p < 0.001) and turning (p < 0.001) tasks.

The hierarchical pattern of sensor performance was preserved across all tasks (resulting in pairwise comparisons of p < 0.001 for each combination of tasks), with lower limb sensors consistently outperforming trunk and upper body sensors. However, the magnitude of performance degradation with increasing task complexity varied by sensor location. This effect was particularly noticeable for the wrist sensor, which showed the steepest drop in performance during stationary and transitional movements compared to other sensor locations (Figure 2).

### Effects of Disease Status

Model performance was robust across both PD and control participants, though performance was significantly higher when assessed on control participants compared to PD participants (p < 0.001), as evaluated using the Friedman test (n = 80 cross validation fold × sensor combinations). While overall performance was only slightly lower in PD participants compared to controls (median F1 scores of 89% and 92%, respectively), the direction of these differences was consistent across all sensor-dataset pairs. Nominally, the impact of disease status varied by sensor location (Figure 4). Lower-limb sensors showed similar performance between groups, demonstrating high stride segmentation accuracy regardless of disease status. However, sensors placed on the head and wrist showed more pronounced performance decreases in PD participants, suggesting that upper body movement patterns associated with PD may impact stride segmentation when using sensors at these locations.

**Figure 4:**
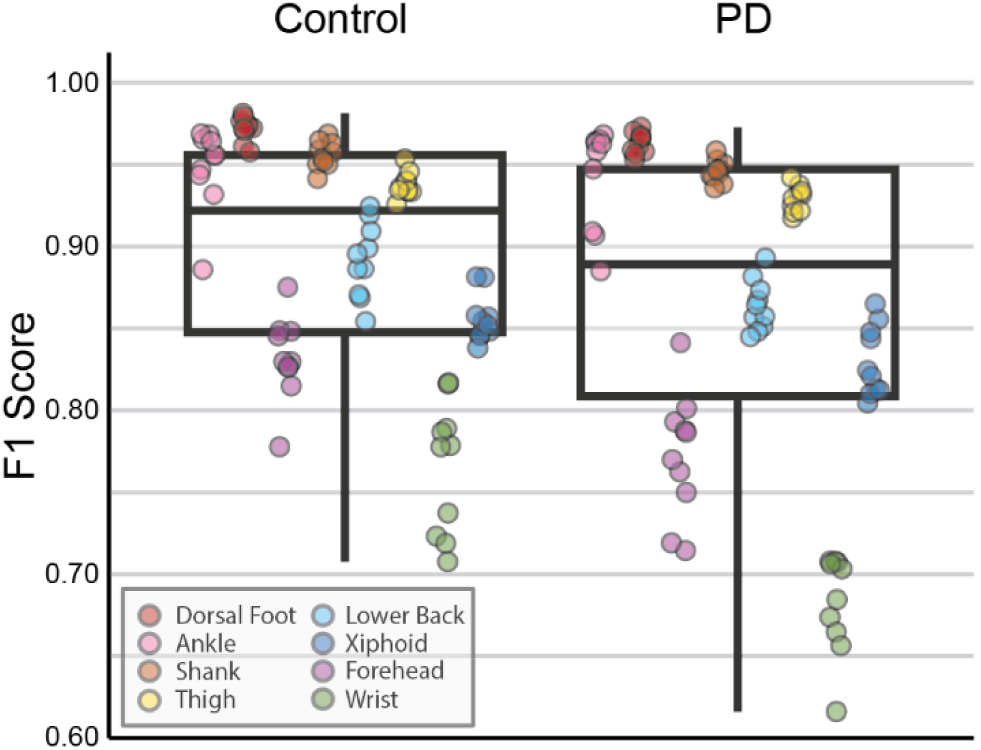
Box and whisker plot comparison of F1 scores between participants with Parkinson’s Disease (PD) compared to control participants (Control). Individual points indicate the F1 score for a particular sensor location as evaluated on the test set of a given cross validation fold when all 81 participants were used for model development.

### Visualizations of Segmented Strides

To visualize the TCN models’ ability to segment strides across diverse sensor locations and movement tasks, we present time-normalized average IMU signals for true positive strides detected from each lower limb (Figure 5) and upper body (Figure 6) sensor. These visualizations used the 81-participant models from the test set of one cross validation fold. Lower-limb sensors exhibited characteristic and consistent waveforms with relatively low variance across strides, particularly during walking. In contrast, signals from the trunk and upper body sensors showed progressively increased variability and less distinct stride-related patterns, particularly during turning. These visualizations align with our quantitative findings, showing how the distance along the kinematic chain affects signal characteristics and, consequently, model performance.

**Figure 5:**
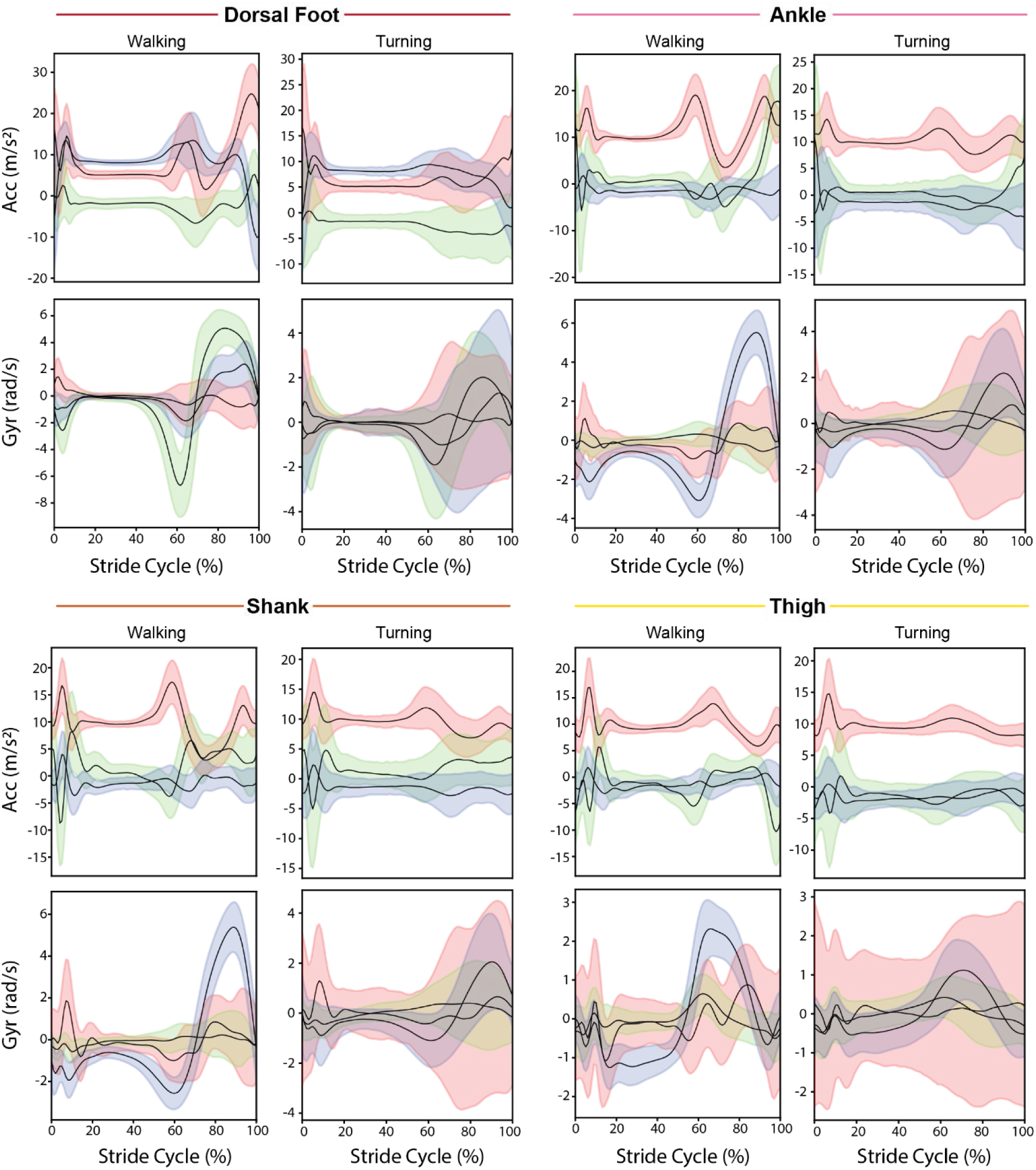
Time-normalized average IMU signals for true positive strides segmented from each lower limb sensor. Black lines show the mean signal for each axis, with colored regions showing ±1 standard deviation (red, blue, and green shadings indicate the X, Y, and Z axes, respectively).

**Figure 6:**
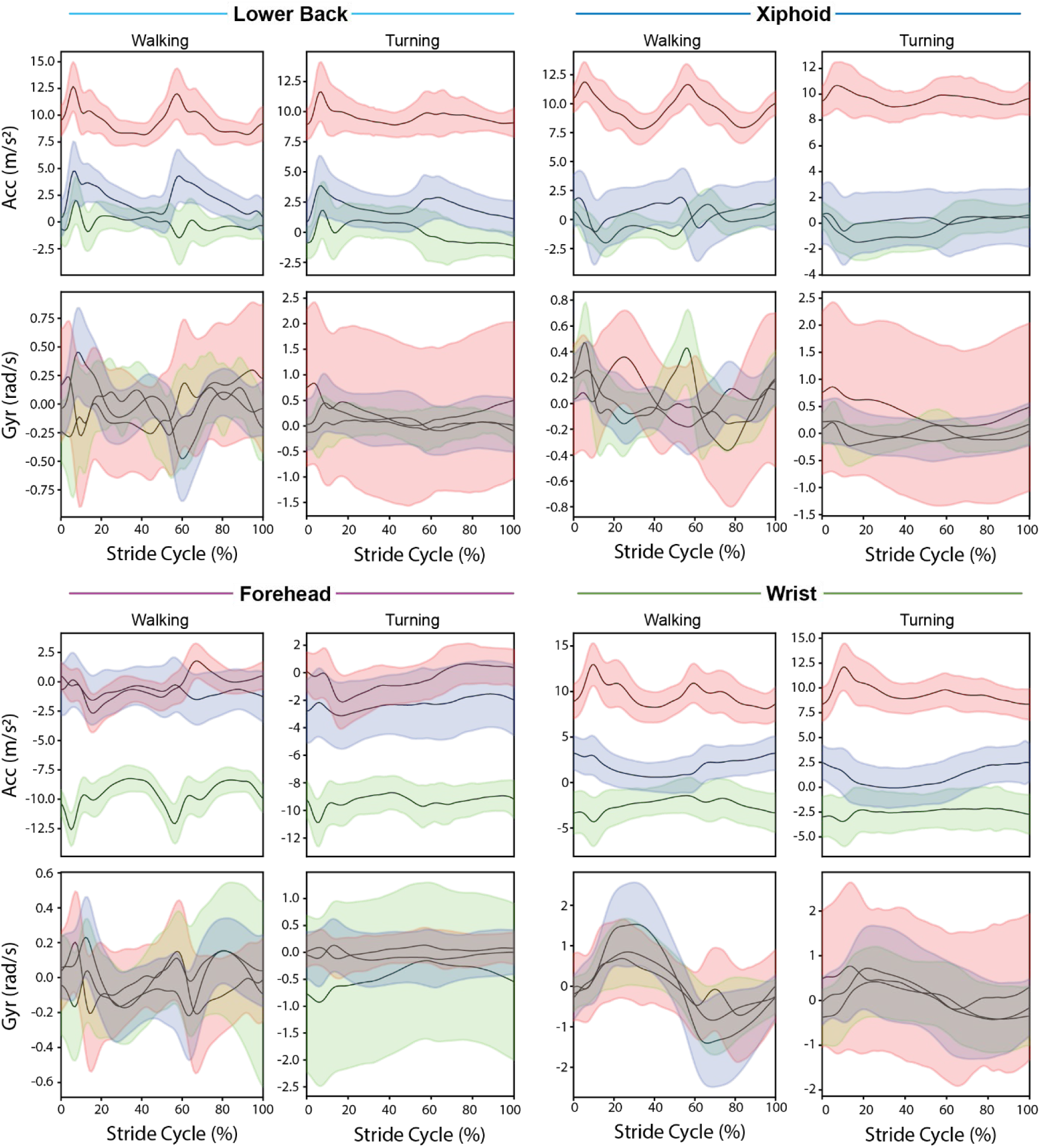
Time-normalized average IMU signals for true positive strides segmented from each upper body sensor. Black lines show the mean signal for each axis, with colored regions showing ±1 standard deviation (red, blue, and green shadings indicate the X, Y, and Z axes, respectively). The relatively lower stride segmentation performance for the upper body sensors should be considered when interpreting these signals.

## Discussion

In this study, we systematically evaluated TCN models for stride segmentation across multiple sensor locations, model development dataset sizes, and tasks. Our results demonstrate that high model performance (F1 scores > 95%) is achievable with lower limb sensors during walking tasks, even with surprisingly small training datasets consisting of 10 participants. However, performance varied substantially by sensor location and declined during complex movements, particularly for sensors placed farther up the kinematic chain from the feet. These findings have important implications for the development of digital health technologies that assess gait in clinical or free-living settings. Most notably, our results indicate that it is important to tailor the evaluation of digital gait assessment tools to the use case, as performance achievable during controlled walking may not translate to more complex movement tasks performed in real-world settings.

Our finding that high stride segmentation performance can be achieved with relatively small model development datasets challenges conventional wisdom about deep learning approaches requiring “big data”. For lower limb sensors during walking tasks, models achieved F1 scores above 90% with only 5-10 participants in the training and validation sets, suggesting that the biomechanical constraints of walking result in relatively consistent IMU signal patterns across participants within our dataset. However, this efficiency in training data requirements did not extend to more complex movement tasks or to sensors placed farther along the kinematic chain from the feet. The wrist sensor, for instance, showed an inability to reach near maximal performance with the complete model development dataset, even under laboratory conditions where there was minimal use of the upper extremities and likely relatively low variability compared to free-living. Additionally, the wrist sensor showed lower performance in the PD group (Figure 4). These results suggest that the data requirements for developing reliable TCN models is highly dependent on movement task complexity and sensor location, with implications for study design and timeline planning in digital health technology development.

The degradation in model performance during complex movements highlights important contextual considerations for stride segmentation algorithms. While many laboratory studies have focused primarily on straight-line or treadmill walking^10,14,18,20^, our analysis systematically evaluated performance across transitions, turns, and other movements, showing substantial declines in performance during these more complex tasks. For instance, F1 scores during turning movements were consistently lower than during walking, even for lower limb sensors. Prior research studies of IMU-based gait assessment in free-living environments have addressed this challenge by limiting analysis to sustained walking bouts, typically identified through rule-based approaches that require multiple consecutive strides over defined time periods^15,16,28^. While this approach effectively captures walking patterns similar to our “walking” task condition, it necessarily excludes more complex movements. Indeed, even the walking task condition modeled here included gait initiation and termination strides, with model performance maintained at high levels. These results suggest that expanding the utility of IMU-based gait assessment to include complex movements, both in the laboratory and free-living environment, remains an important area for future development.

The drop off of stride segmentation performance as sensor location moves up the body should be considered in light of user experience for applications including clinical assessments and remote monitoring. For one-time clinical gait assessments, placing sensors on the feet or lower limbs may not be burdensome and would provide the most reliable measurements. However, for continuous remote monitoring over longer periods of time, user experience and compliance become critical factors^29–31^. Sensors on the midline or wrist may become more attractive^32^, especially considering the widespread adoption of smartwatches and the existing infrastructure that comes with them. Yet our results show substantially lower performance for wrist-mounted sensors across all tasks, with F1 scores nearly 50% lower than foot sensors during complex movements. Furthermore, the diversity of wrist movements recorded in our study was limited relative to those present in daily life. This performance gap, combined with the reliability of foot and ankle-mounted sensors, suggests an opportunity where lower limb sensors could serve as reference devices for characterizing and improving algorithms developed for wrist-worn devices.

Our study extends previous work on deep learning-based stride segmentation algorithms in several ways. While earlier studies primarily focused on algorithmic development and validation during walking^16,19,20^, we present an assessment of TCN performance across multiple movement tasks and sensor locations. Our results align with previous findings that algorithms which learn directly from data show promise for handling complex tasks with greater movement variability^13^. However, our findings suggest that accurately segmenting strides during complex movements remains challenging across algorithmic approaches, though these limitations could potentially be overcome through the development of high-quality training datasets that better represent complex movements.

When evaluating stride segmentation performance, particularly at the high end of reported F1 scores, it is important to consider fundamental challenges in establishing ground truth. While initial contacts during walking are well-defined biomechanical events that can be reliably detected with instrumented walkways^18^, pressure-sensing insoles^16^, or optical motion capture systems^20^, the boundaries between strides become less clearly identifiable during complex movements or when gait is altered by pathology. For example, during 180-degree turns, participants may drag their feet, making it unclear exactly when a new stride begins. Current approaches to establishing ground truth typically combine specialized hardware, automated labeling algorithms, and human judgement to review and correct imperfectly placed labels, introducing an inherent level of subjectivity in model training and evaluation. This challenge of label noise may help explain why even our best-performing models plateau below perfect accuracies. Studies also vary in how they define successful stride detection, with different time windows used to identify true positives, ranging from ±16 milliseconds^24^ to ±250 milliseconds^16^. These methodological differences, while appropriate for different applications, should be considered when comparing performance metrics across studies.

Overall, our findings suggest that it is important to tailor performance testing for digital health technologies using stride segmentation algorithms to their clinical application. For example, algorithms for applications focused on clinical assessments of walking in controlled environments may be trained and characterized with less extensive datasets, particularly when using lower limb sensors, compared to those aimed at continuous monitoring of diverse movements.

Several limitations in our study should be considered. First, all data were collected in a controlled laboratory environment with specific protocols and standardized walking paths on level ground. Although we included complex movements like turns and transitions, these were still more constrained than mobility in daily life. Second, our analysis focused on people with Parkinson’s disease and healthy controls. Validation in other populations with altered gait patterns will be needed for broader applications, especially in impairments that lead to more heterogeneous gait patterns (e.g., cerebral palsy, stroke). The development of open-source, population-specific datasets would enable more targeted algorithm development for different clinical populations. Lastly, ‘strides’ are typically well-defined during walking, but in this study, we also identified initial contact events (i.e., stride boundaries) in complex movements like 180-degree turns and transitions into and out of chairs. While these movements do not constitute strides in the traditional walking sense, they likely contain relevant health information.

Future work should focus on evaluating and extending these approaches to free-living environments, where movement patterns are more diverse and ground truth measurements are more challenging to obtain. This will require the development and refinement of novel ground truth systems suitable for home-based data collection, potentially leveraging pressure-sensing insoles^33^ or video-based motion capture^34^. Finally, establishing appropriate performance requirements for stride segmentation algorithms remains an open challenge, as these algorithms typically serve as sub-modules within larger gait assessment pipelines, and the necessary F1 scores will ultimately depend on the clinical application, how errors in identified stride boundaries propagate to downstream metrics, and minimally important differences in derived gait metrics. Our findings demonstrate that TCNs can achieve robust stride segmentation performance across varied mobility tasks, particularly when using lower limb sensors, representing an important advancement in wearables-based gait assessment. Given that accurate stride segmentation serves as the foundation for deriving meaningful gait metrics, these results support the continued development of TCN-based approaches for digital gait assessment in both clinical and remote settings.

## Methods

### Dataset and Data Collection

We analyzed data from the open-access WearGait-PD dataset^27,35^ containing movement data from older adults with and without PD performing standardized gait tasks in a laboratory setting. Data collection was performed at three clinical sites (one in Seattle, WA and two in Baltimore, MD) using a standardized protocol that included straight-line walking, 180-degree turns, standing, and other movement tasks. Movement data was captured using Movella/Xsens MTw Awinda inertial measurement units (IMUs) placed at eight body locations: mid-forehead, sternum, lower-back (L4/L5), wrists, lateral thighs, lateral shanks, lateral ankles (proximal to malleoli), and dorsal feet. Sensors on the limbs (e.g., ankles, wrists, etc.) were worn bilaterally. A ProtoKinetics Zeno instrumented walkway system provided ground truth initial contact detection through embedded pressure sensors. The IMU and walkway systems were time synchronized with a TTL pulse, and both systems sampled at 100 Hz.

Participants completed experimental conditions designed to capture diverse movement tasks. These included four passes across the walkway of self-paced and hurried-pace walking, with turns occurring both on and off the walkway, and three repetitions of the Timed Up and Go (TUG) test, with turns completed on the walkway. Participants completed an additional self-paced walking task, with 180-degree turns occurring on the walkway, where turning direction was controlled such that two left turns and two right turns were recorded. Video recordings enabled frame-by-frame annotations of specific movement tasks (e.g., ‘sitting’, ‘walking’, ‘turning’) within each experimental condition. Complete details of the study protocol, including task definitions and annotation procedures, are available in the online data repository and associated publication^27,35^.

### Data Processing and Quality Control

IMU data preprocessing included gap-filling of missing data using cubic splines, with forward/backward filling applied at recording boundaries. Acceleration and gyroscope signals were low-pass filtered using a 4^th^-order, zero-lag Butterworth filter with a −3 dB cutoff frequency of 20 Hz. For sensors on the arms and legs, left limb sensor axes were transformed to align with right limb sensors to ensure consistent interpretation of movement directions relative to the body midline.

Data quality screening was performed to identify and remove regions with wireless signal dropout or initial contact events occurring off the instrumented walkway. This automated process, detailed in the Supplementary Materials, evaluated both IMU data quality (identifying regions of missing data) and walkway data quality (identifying periods when participants stepped off the active sensing area). Only data segments meeting both quality criteria were retained for analysis.

Initial contact events were identified in the walkway data by applying a rising-edge detection algorithm to the foot-ground contact Boolean signal for each leg automatically provided by the instrumented walkway. This operation resulted in a rising-edge pulse waveform for each leg, where frames containing an initial contact event contain 1’s and all other frames contain zeros. For model training, these binary pulses were convolved with a triangular kernel to create probability score waveforms. This approach allowed for continuous probability estimates of initial contact events rather than binary classification. During model evaluation, the original rising-edge pulses were used to define event timing.

### Deep Learning Framework

We developed Temporal Convolution Network (TCN) models to detect initial contact events from IMU data. The models transform IMU acceleration and gyroscope signals (6 channels × T time steps) to probability scores between zero and one (1 channel × T time steps), where higher values indicate greater likelihood of an initial contact event. A peak-finding algorithm with a magnitude threshold is then applied to these probability scores to identify specific initial contact time indices. The TCN architecture consists of dilated convolutions arranged in residual blocks, allowing the model to capture long-range temporal dependencies while maintaining computation efficiency (Figure 7).

**Figure 7:**
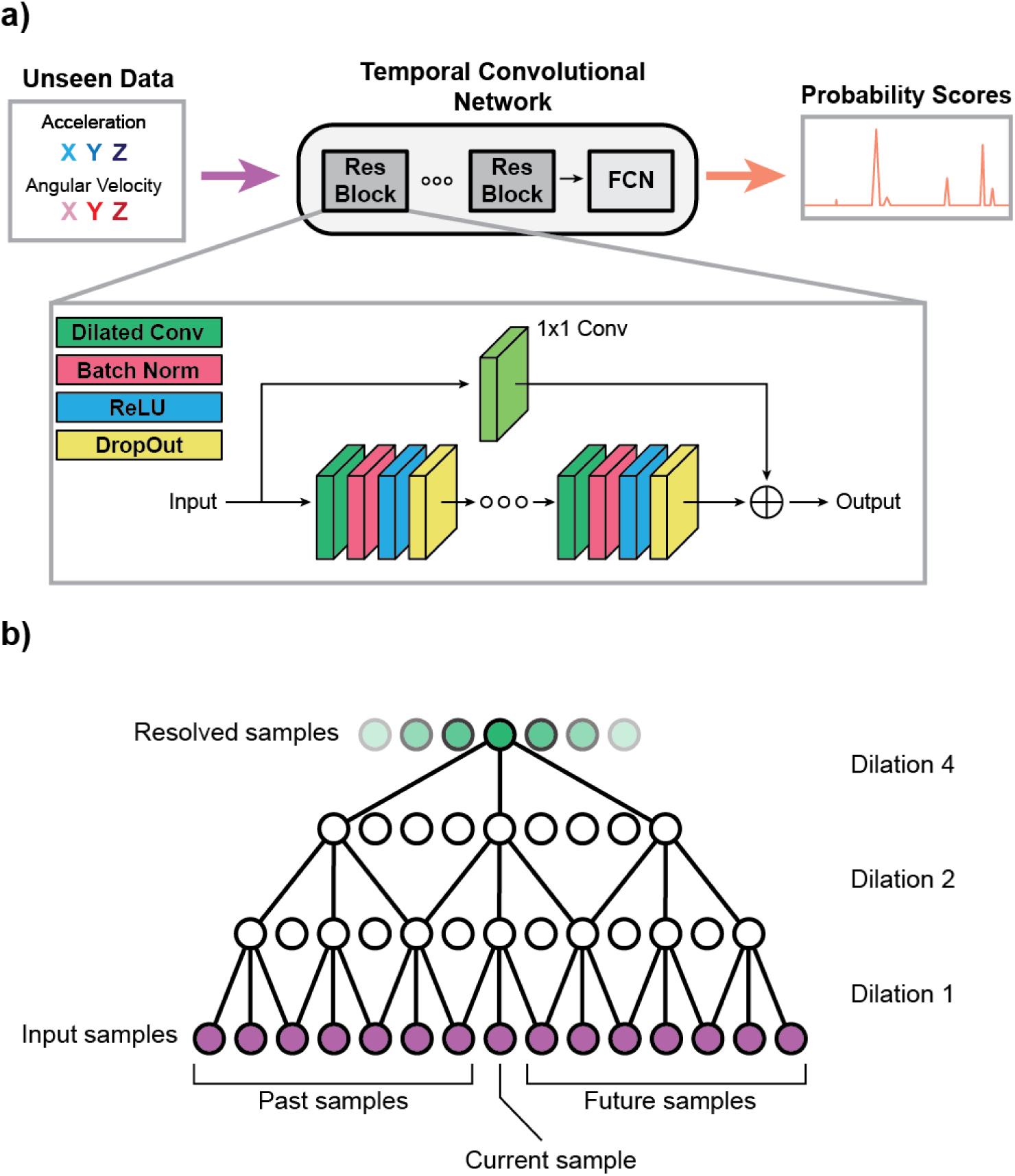
Representative model architecture for our deep learning framework. a) A high-level representation of how our model architecture is used to process three acceleration and three gyroscope signals for a given sensor and produce probability scores. The inset depicts a more detailed representation of one residual block, which contains diluted convolutional layers, batch norm layers, rectified linear unit (ReLU) layers, and drop out layers. The ellipses between the residual blocks and within the inset indicate that the number of residual blocks and number of layers were left open as a hyperparameter. b) A high-level representation of a set of diluted convolutional, batch norm, ReLU, and dropout layers, in which information from past and future samples are synthesized through several dilutions. In practice, the number of levels of dilution were left open as a hyperparameter.

During training, input signals were normalized channel-wise to have zero mean and unit variance. To prevent amplification of noise in low-magnitude signals (e.g., during sitting), normalization was only applied when it reduced the maximum absolute value in the waveform. Models were trained using the Adam optimizer with mean squared error loss and an early stopping mechanism that ceased training when validation loss stopped decreasing across 4 consecutive epochs.

Model hyperparameters, including network architecture parameters and training settings, were optimized independently for each unique combination of sensor location, training data quantity, and cross validation fold using Optuna^36^. Complete details of the hyperparameter optimization process are provided in the Supplementary Materials. All TCN models were developed using Keras and TensorFlow, specifically with the Keras-TCN package^37^. Model training was conducted on a Dell Precision 7780 laptop with an NVIDIA RTX 5000 GPU.

### Cross Validation Strategy

We employed a 10-fold cross validation framework to assess model generalizability, with each fold maintaining distinct training, validation, and test sets. For each fold, the test set consistently included 40 participants, while the remaining participants were split evenly between training and validation sets. The allocation of participants to sets was stratified based on sex, disease state (PD vs control), and Hoehn and Yahr score (for PD participants) to ensure balanced representation across sets.

To evaluate the impact of training data quantity, we systematically varied the number of participants included in the training and validation sets (from 2 to 81 total participants split equally between sets) while maintaining the consistent 40-participant test set. Within each fold, participants were added sequentially such that any dataset of size *n* was a subset of all larger datasets. Models were developed for development dataset (training plus validation) sizes of 2, 4, 6, 8, 10, 12, 14, 16, 18, 20, 30, 40, 60, and 81 participants. This approach was repeated independently for each sensor location, resulting in the development of unique models for each combination of sensor location, training data quantity, and cross validation fold.

In cases where a participant was known to be missing all data for a given sensor due to equipment malfunctions, they were always placed in the validation set, occasionally resulting in 39-40 participants with usable data in the validation set for certain sensor locations. In the case of using all 81 participants, the validation dataset nominally contained 41 participants.

### Model Evaluation and Statistics

Model performance was evaluated by comparing algorithm-detected initial contact events to ground truth events from the instrumented walkway. True positives (TP) were defined as algorithm-detected events within ±30 milliseconds of walkway-labeled events (three samples on either side of the ground truth), with only one TP counted per true event. False Positives (FP) were either algorithm-detected events outside this window or additional detections beyond the closest TP for a given walkway event. False negatives (FN) were walkway-detected events missed by the algorithm.

Using the labeled data points, we calculated precision, recall, and F1 score to quantify the algorithm’s performance. Precision, representing the proportion of correctly identified events among all events detected by the algorithm, was computed as:

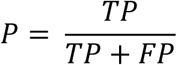

Recall, indicating the proportion of actual events correctly identified by the algorithm as compared to the walkway, was computed as:

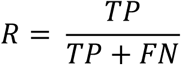

The F1 score was computed as the harmonic mean of precision and recall:

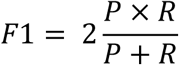

An F1 score close to 1.0 indicates that the model captures nearly all relevant events (high recall) and reporting a low number of false positives (high precision).

To assess performance across different movement tasks across our experimental conditions, we leveraged the video-based task annotations to analyze three distinct categories of movement task: (1) walking tasks, which included gait initiation, straight-line walking, and gait termination, (2) 180-degree turns, and (3) stationary and transitional movements, which included the labels ‘sitting’, ‘standing’, ‘sit-to-stand’, and ‘turn-to-sit’. Model performance metrics were computed independently for each movement task category. Additionally, we compared model performance between PD and control participants to evaluate the robustness of our approach across both groups.

Mean F1 score was computed across cross validation folds, and 95% confidence intervals were computed using a bootstrapping approach with 100,000 random samples with replacement. Statistical comparisons differed based on research question. Differences in performance between different model development data quantities were evaluated using descriptive statistics alone. Omnibus comparisons to evaluate differences in performance across sensor locations, movement tasks, and disease states were performed using the non-parametric Friedman test, as the F1 scores are not normally distributed (primarily due to a ceiling effect) and the 10-fold cross validation resulted in participants shared across datasets, resulting in non-independent samples. For pairwise comparisons of performance across sensors and tasks, we used a post-hoc Nemenyi test as it is also non-parametric, and controls for family-wise error rate. All statistical tests were two-tailed with an alpha value of 0.05. Statistics were computed using RStudio (Posit, Boston, MA, Version 2023.09.0 Build 463).

As a final descriptive analysis, we selected representative strides identified by the algorithm from the walking and turning tasks to demonstrate the breadth of different types of movements this approach is capable of segmenting. To segment strides, we extracted regions of IMU data between two adjacent true positive initial contact events for all participants in the test set of one cross validation fold.

## Code Availability

The underlying code for this study is not publicly available but may be made available to qualified researchers on reasonable request from the corresponding author.

## Data Availability

The raw data analyzed during the current study are available in the "Wearables for gait in Parkinson's Disease and age-matched controls (WearGait-PD)" repository, https://doi.org/10.7303/syn52540892.

https://www.synapse.org/Synapse:syn52540892/wiki/623751

## Acknowledgements

This study was funded by CDRH’s Critical Path Initiative (CP2024-1275) as well as the Veterans Affairs Rehabilitation Research and Development Seed Award (A2357C) and VA Puget Sound Health Care System Seed Award (SMUI). This project was supported in part by an appointment to the Research Participation Program at the Center for Devices and Radiological Health, U.S. Food and Drug Administration, administered by the Oak Ridge Institute for Science and Education through an interagency agreement between the U.S. Department of Energy and FDA. No funder played any role in the study design, data collection, analysis, interpretation of the data, or writing of this manuscript.

## Author Contributions

Conceptualization: AA, MG, BM, SH, VK, KK; Methodology: AA, MG; Software: AA, MG; Validation: AA; Formal Analysis: AA, MG; Investigation: AA, MG, DE, NK, IZ, SH, BM, KK; Data Curation: AA, MG, DE, NK, IZ, SH, KK; Writing – Original Draft: AA; Writing – Review & Editing: MG, BM, SH, KK, VK, NK, IZ, DE. Visualization: AA, MG; Funding Acquisition: KK, AA, BM. Contribution categories were drawn from the CRediT (Contributor Roles Taxonomy) framework. All authors read and approved the final version of the manuscript.

## Disclaimer

The mention of commercial products, their sources, or their use in connection with material reported herein is not to be construed as either an actual or implied endorsement of such products by the Department of Health and Human Services.

