## Supplementary Materials for "Deep learning-based stride segmentation with wearable sensors: effects of data quantity, sensor location, and task"

**Affiliations**


#### Data Quantity Breakdown Across Sensor Locations

Data quantity, in terms of both time and gait events, varied by sensor location due to wireless signal dropout. We quantified the number of initial contact events detected by the walkway in and the total duration of recordings in the open-access dataset after our signal quality screening algorithm was applied. The results are shown in Table S1 below:

Table S1: Data quantity across the entire open-access dataset after the signal quality algorithm was applied.

| ***Sensor*** | ***Total Events*** | ***Duration (min)*** | ***Mean Strides Per Participant*** | ***SD Strides Per Participant*** | ***Total Participants*** |
|---|---|---|---|---|---|
| *Dorsal Foot* | 26,418 | 342 | 220 | 49 | 120 |
| *Ankle* | 26,701 | 346 | 223 | 51 | 120 |
| *Shank* | 26,691 | 344 | 222 | 50 | 120 |
| *Thigh* | 26,407 | 342 | 222 | 53 | 119 |
| *Lower Back* | 26,910 | 348 | 222 | 51 | 121 |
| *Xiphoid* | 27,018 | 350 | 223 | 52 | 121 |
| *Forehead* | 26,679 | 346 | 222 | 51 | 120 |
| *Wrist* | 26,816 | 347 | 222 | 50 | 121 |
| ***Mean*** | 26,663 | 346 | 222 | 51 | 120 |

### Statistical Comparisons of Model Performance Across Sensor Locations

Performance was compared across sensor locations for the 81-participant models. First, the non-parametric Friedman test was used to determine that significant differences existed across groups. Then, a post-hoc Nemenyi test was used to make pairwise comparisons between performance. Table S2 shows the p-values resulting from the statistical comparisons:

Table S2: Post-hoc Nemenyi test results (p-values) for pairwise comparisons of average F1 score across cross validation folds for each sensor location. Each comparison was performed when 81 participants were used for model development. Bold entries indicate statistically significant differences.

| | *Ankle* | *Dorsal Foot* | *Forehead* | *Shank* | *Lower Back* | *Thigh* | *Wrist* |
|---|---|---|---|---|---|---|---|
| *Dorsal Foot* | 0.779 | - | - | - | - | - | - |
| *Forehead* | **0.003** | **<0.001** | - | - | - | - | - |
| *Shank* | 1.000 | 0.871 | **0.002** | - | - | - | - |
| *Lower Back* | 0.357 | **0.005** | 0.724 | 0.254 | - | - | - |
| *Thigh* | 0.974 | 0.172 | 0.088 | 0.936 | 0.936 | - | - |
| *Wrist* | **<0.001** | **<0.001** | 0.974 | **<0.001** | 0.139 | **0.003** | - |
| *Xiphoid* | 0.053 | **<0.001** | 0.992 | **0.030** | 0.992 | 0.476 | 0.602 |

### Details on Signal Quality Screening Algorithm

A signal quality screening algorithm was developed to address two primary sources of poor data quality: missing IMU data due to wireless signal dropout and heel strike events occurring off of the instrumented walkway. In the latter scenario, there is no ground truth heel strike event to train on, which introduces label noise. While some off-walkway steps were intentional in tasks starting and ending off the walkway (self-paced and hurried pace walking), others were accidental missteps in tasks intended to be confined to the walkway.

The algorithm, applied to all files across all participants prior to model development, consisted of the following steps:

1. <u>IMU Quality Assessment:</u> A Boolean "IMU Quality" signal was created, set to False during regions with 7 or more consecutive missing frames in either the accelerometer or gyroscope signals, and set to True elsewhere. This approach allows for splining of smaller gaps while flagging regions with substantial data loss.
2. <u>Walkway Quality Assessment:</u> Using the walkway's built-in Boolean time series signals for left and right foot-ground contact, we created an 'Either Foot-Ground

Contact' waveform using the OR operation. A "Walkway Quality" signal was then derived by removing a 50-frame (0.5 second) buffer from the beginning and end of each True region in the 'Either Foot-Ground Contact' waveform. This buffer helps eliminate heel strike events that occurred off the walkway while one foot was still in contact with the walkway.

3. Total Quality Assessment: A "Total Quality" signal was generated using the AND operation on both the 'IMU Quality' and 'Walkway Quality' signals, identifying high-quality data regions within each file.

This screening process ensured that high-quality data from the IMUs and walkway were used in subsequent analyses, minimizing the impact of signal dropout and off-walkway events on model development and evaluation. In order to avoid data leakage, parameters in the quality screening algorithm were not tuned to maximize downstream model performance.

Figure S1 shows a representative example of an instance where the quality screening algorithm automatically removed a region of low-quality data.

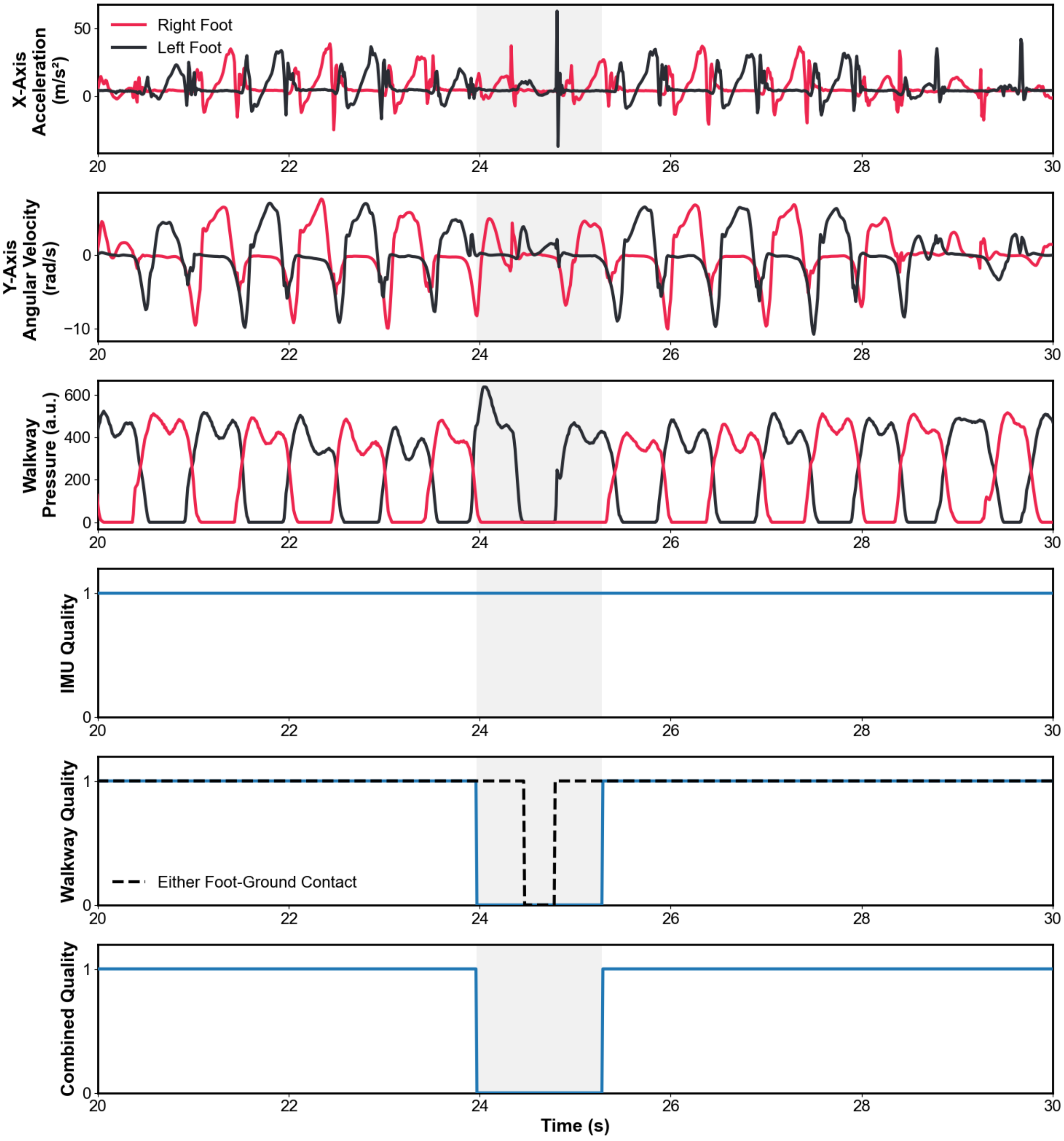


Figure S1: An example where the data quality screening algorithm removed a section of a file (grey region) when the participant accidentally stepped off of the instrumented walkway during a 180-degree turn. Acceleration and angular velocity signals from the dorsal foot sensors are shown on the top two plots, as well as relative pressure waveforms from the walkway (third panel). By inspection, it can be determined that the right foot did not contact the walkway during the low-quality region. The bottom three panels show Boolean signals used to determine if the data is high or low quality.

#### Model Architecture and Hyperparameter Optimization

The Temporal Convolutional Network (TCN) architecture consists of dilated convolutional layers arranged in residual blocks with layer normalization, weight normalization, and skip connections. Each model takes as input six channels of IMU data (three accelerometer and gyroscope channels) and outputs a single-channel probability score between 0 and 1 indicating the likelihood of a heel strike event at each time point. Our implementation uses non-causal convolutions, allowing the model to use both past and future IMU signals when determining the probability of a heel strike at any given time point.

For sensors placed on limbs, a single model was capable of detecting left and right heel strikes due to the harmonized axes. For midline sensors (lower back, xiphoid, and forehead), two models were developed per location to detect heel strikes on each side of the body. Model performance was averaged between left and right sides for these midline sensors in all analyses.

During training, input data was processed in fixed-length windows with 50% overlap. Each window underwent a conditional normalization process – if normalizing all six IMU channels to zero mean and unit variance would reduce the maximum absolute value across all channels in the window, then normalization was applied. This approach prevented amplification of noise in low-magnitude signals, which as during sitting or standing. Target data for training was generated by convolving the binary heel strike indicators from the instrumented walkway with a symmetric triangular kernel spanning 160 ms (80 ms on each side of the event), creating continuous probability scores rather than binary classifications.

Models were trained using the Adam optimizer with a fixed batch size of 32 and mean squared error loss function. Training proceeded for a maximum of 500 epochs with an early stopping mechanism that terminated training if validation loss did not improve for 4 consecutive epochs. This configuration was maintained across all model development scenarios.

Hyperparameter optimization was performed using Optuna's Tree-structured Parzen Estimators (TPE) algorithm to explore the model architecture and training parameters shown in Table S3.

Table S3: Hyperparameters and ranges searched for each sensor, data quantity, and cross validation folder. All parameters were searched as integers, except for learning rate and dropout rate, which were floats.

| ***Hyperparameter*** | ***Search Range*** |
|---|---|
| *Residual Blocks* | 1, 2, 3 |
| *Layers Per Block* | 1, 2, …, 6, 7 |
| *Filters Per Layer* | 10, 11, …, 49, 50 |
| *Kernel Size* | 5, 6, …, 11, 12 |
| *Window Length* | 200, 300, 400, 500 |
| *Learning Rate* | 1e-3 – 1e-2 |
| *Dropout Rate* | 0.05 – 0.4 |

Eight hyperparameter combinations were evaluated for each unique combination of sensor location (8), model development dataset size (14), and cross-validation fold (10), resulting in 8,960 total models trained during optimization. For each combination, the model achieving the highest F1 score on the validation dataset was selected.

On some occasions, particularly with training runs using small numbers of participants in the training dataset, the optimal model architecture selected by Optuna failed to successfully train (i.e., validation loss did not decrease between the first and last training epoch). On these occasions, the hyperparameter search process was restarted with 50 iterations rather than 8.

Following model training, heel strike events were identified by applying a peak-finding algorithm to the output probability scores with a minimum distance constraint of 25 samples between adjacent peaks. The probability threshold for peak detection was optimized by evaluating F1 scores on the validation dataset across thresholds between 0 and 1, selecting the threshold that maximized performance.
